# Sex Differences Across the Donor Heart Offer-to-Outcome Pathway: A Linked National Cohort Study

**DOI:** 10.64898/2026.10.21.26366181

**Authors:** J. Sam Meyer, Olivia Gilbert, Adrian L. Lata, Eliana Pickholz, Madhav Swaminathan, Edward H. Kincaid, Yaron D. Barac

## Abstract

**Background:** Sex differences occur across heart transplantation, but where they become apparent between donor-heart offers and post-transplant outcomes is uncertain.

**Methods:** We linked UNOS STAR Q2 2025 candidate, donor, transplant, and PTR match-run data across pre- and post-2018 eras. Analyses included 51,970 listing episodes, 1,388,553 actual offers, and 38,417 recipients and evaluated access, conditional final acceptance, donor profiles, severe predicted heart mass (PHM) undersizing (ratio <0.86), treated rejection, and 365-day survival.

**Results:** Transplant access increased in both sexes without evidence of sex-by-era modification (P=.133). Offers made to female candidates had higher adjusted conditional final-acceptance probabilities in both eras (interaction P=.172). After recipient-size standardization, severe PHM undersizing among realized offers remained less frequent for women than men pre-policy (28.9% vs 38.3%) and post-policy (25.5% vs 35.4%); acceptance of undersized offers was not lower for female candidates (three-way interaction P=.800). The threshold donor composite showed modest differences that reversed direction across eras. Adjusted treated rejection remained higher among women (male vs female OR, 0.667; 95% CI, 0.589-0.756), including after post-policy cPRA adjustment (OR, 0.678; 95% CI, 0.583-0.790). Adjusted post-policy 365-day survival was 91.0% in women and 92.3% in men; the sex difference did not change across eras (−0.64 percentage points; 95% CI, −1.97 to 0.68; P=.341).

**Conclusions:** Sex-associated patterns were stage-specific and multidirectional. The final-graft PHM difference was already evident among realized offers after size standardization and was not explained solely by lower acceptance of undersized offers for female candidates. Women nonetheless had more treated rejection, while short-term survival differences did not change significantly across eras. Transplantation performance should be assessed separately across offer exposure, acceptance, matching, and outcomes.

**What Is Known:**

- Sex differences have been reported in heart-transplant access, donor-heart acceptance, sensitization, rejection, and survival, but these stages have usually been examined separately.
- Offer exposure, conditional acceptance, donor-recipient size matching, and downstream outcomes may not show the same direction of association with recipient sex.

**What the Study Adds:**

- By linking 1.39 million actual donor-heart offers to listing and transplant records, this study found higher adjusted conditional final-acceptance probabilities for offers made to female candidates in both allocation eras.
- After recipient-size standardization, women encountered and received fewer severely PHM-undersized hearts; the difference was already apparent among realized offers and was not explained solely by lower acceptance of undersized offers for female candidates.
- Conventional donor profiles, treated rejection, and survival did not move uniformly: women had more adjusted registry-reported treated rejection, whereas short-term survival differences did not change significantly across eras.

## Introduction

Women comprise approximately one-quarter of adult heart transplant recipients, yet sex-associated differences persist across referral, mechanical circulatory support, waitlist outcomes, donor selection, sensitization, rejection, and survival. (1–5) The 2018 U.S. adult heart allocation revision introduced more granular urgency tiers and broader geographic sharing, changing transplantation access, procurement patterns, and center practice. (6–10) These changes provide an important policy context in which to examine where along the transplant pathway sex differences become apparent.

DeFilippis et al. reported greater transplantation opportunities for women during the first 2 years after the allocation revision; Purohit et al. compared sex-specific waitlist and post-transplant outcomes under the previous and revised systems; and Ramu et al. characterized 5-year outcomes under the contemporary policy. (11–13) Using post-2018 match-run data, Breathett et al. found higher adjusted conditional donor-heart acceptance for offers made to women. (14) However, no national study has linked actual offer exposure and conditional acceptance to final donor-recipient matching, utilized donor profiles, treated rejection, and survival within the same cohort.

Donor-recipient size matching is central to this question because historical signals attributed to donor-recipient sex mismatch are strongly intertwined with cardiac size. (15,16) Kransdorf et al. established PHM as a clinically informative matching metric and identified increased 1-year mortality with severe undersizing, although subsequent studies suggest that its prognostic importance varies by recipient phenotype and allocation era. (17–19) Conventional donor characteristics are also relevant but heterogeneous and context dependent. (20) Sensitization, rejection, and survival provide complementary downstream endpoints, and demographic associations may differ across these outcomes. (4,21–27)

We therefore linked UNOS STAR and PTR match-run data to evaluate sex-associated patterns from listing and donor-heart offers through transplantation and early outcomes. The principal stage-localization analysis examined where differences in PHM matching were detectable across realized offer exposure, conditional final acceptance, and final transplantation after accounting for recipient size. Transplant access, conventional donor profiles, registry-reported treated rejection, and survival were evaluated as complementary domains, with interaction terms assessing whether sex associations differed across allocation eras.

## Methods

### Data Sources and Study Design

We conducted a retrospective national cohort study using the UNOS STAR Q2 2025 release, with follow-up through June 30, 2025, linked to heart and heart-lung PTR match-run data. Three complementary analytic populations were assembled: (1) adult first-time isolated-heart listing episodes for waitlist and first-offer analyses; (2) candidate-level PTR actual offers for offer-exposure and conditional-acceptance analyses; and (3 adult first-time isolated-heart transplant recipients for final donor profiles, PHM matching, treated rejection, and survival. Data were deidentified, and the institutional review board determined the study exempt (IRB No. 172, Pro00073879; May 29, 2016); informed consent was waived. Reporting followed STROBE recommendations.

Analyses compared the pre-policy era (June 21, 2012-October 17, 2018) with the post-policy era (October 18, 2018-June 30, 2025). We excluded candidates and recipients undergoing retransplantation, multiorgan transplantation, heart-lung transplantation, or non-heart thoracic transplantation. Recipient sex was analyzed as female or male as recorded in UNOS; gender identity is not captured. The listing cohort comprised 51,970 qualifying episodes among 49,569 unique patients. The transplant cohort comprised 38,417 recipients: 3,959 women and 11,251 men pre-policy and 6,159 women and 17,048 men post-policy.

### Donor-Heart Offer Cohort and Outcomes

PTR records were restricted to heart offers. An actual offer was defined by OFFER_ACCEPT values N, Y, or Z; bypass records (B) were excluded. Final acceptance was defined as Y. Provisional acceptance (Z) was treated as nonacceptance in the primary model and excluded in a sensitivity analysis. We identified 1,388,553 actual offers during the study period. Candidates were linked to the listing cohort by waitlist identifier, and offer donors were linked to the deceased-donor file by donor identifier. Among transplant recipients, at least one preceding actual offer was captured for 93.1% pre-policy and 95.8% post-policy. Offer-level PHM was calculable for 86.1% of actual offers overall and at least 83.5% of each sex-by-era group.

Time to first actual offer was measured from listing, with waitlist death or removal for clinical deterioration treated as competing events. Conditional final acceptance was modeled per offer among listing episodes still under observation and without an earlier final acceptance; records were retained through the first final acceptance, and the primary analysis was limited to the first 50 offers. The binomial-logit model included recipient sex, era, their interaction, a natural spline for offer number (4 df), sex- and era-by-offer-number interactions, recipient age, race/ethnicity, blood type, diagnosis, BMI, log match sequence, and listing-center fixed effects. Standard errors were clustered by listing episode and donor. A donor-adjusted sensitivity additionally included donor age, sex, LVEF, diabetes, cardiac arrest/CPR, and offer-level PHM ratio. Detailed coding and linkage quality assessments are provided in the Supplemental Material.

### Predicted Heart Mass and Size Matching

Predicted left- and right-ventricular masses were calculated from established sex-specific equations incorporating age, height, and weight; PHM was their sum. (17) Donor-recipient PHM ratio was calculated as donor PHM divided by recipient PHM. Severe undersizing was defined as a ratio <0.86. PHM was evaluated for each actual offer and for the ultimately transplanted graft.

Because recipient sex and body size are closely related, the primary offer-exposure analysis standardized recipient size. Severe PHM undersizing among actual offers was modeled as a function of recipient sex, allocation era, their interaction, a flexible spline for recipient PHM, blood type, primary diagnosis, match sequence/rank, and center fixed effects, with two-way clustered standard errors by candidate and donor. Standardized probabilities placed women and men on the same pooled recipient-PHM and measured covariate distribution. A common-support analysis was restricted to the overlap of the female and male 5th-95th percentile recipient-PHM ranges. Offered donor PHM was also analyzed continuously after adjustment for recipient PHM and the same core covariates. Conditional final acceptance of undersized versus nonundersized offers was assessed with a sex-by-era-by-undersizing interaction.

### Donor Profiles and Post-Transplant Outcomes

Conventional donor and transplant-logistics variables included donor age, LVEF, diabetes, cardiac arrest/CPR, ischemic time, and substance history. As a secondary descriptive summary, we defined a threshold donor-characteristic/logistics composite as the presence of any of the following: donor age >=55 years, LVEF <50%, ischemic time >=4 hours, donor diabetes, or donor cardiac arrest/CPR. Because these components are heterogeneous and context dependent, the composite was not interpreted as a unitary measure of graft quality; alternative definitions were examined in sensitivity analyses.

Registry-reported treated rejection within 1 year was evaluated using the calculated STAR TRTREJ1Y endpoint among recipients transplanted on or before June 30, 2024. Unknown and missing values were excluded rather than classified as rejection or nonrejection. Exact rejection dates were unavailable; therefore, the endpoint was analyzed as a fixed-window binary outcome. The mixed-effects model included recipient sex, allocation era, their interaction, recipient age, race/ethnicity, BMI, primary diagnosis, ECMO, intra-aortic balloon pump, durable VAD/TAH support, HLA mismatch, ABO matching, donor age, donor sex, ischemic time, continuous PHM ratio, and a transplant-center random intercept. For contemporary post-policy recipients, pretransplant cPRA was reconstructed as the most recent valid waitlist-history SENDATACPRA value on or before transplantation and added in a sensitivity model. Pre-policy cPRA was insufficiently complete for valid era-comparative adjustment.

Post-transplant survival was described using Kaplan-Meier estimates. Adjusted 365-day survival was evaluated among recipients transplanted on or before June 30, 2024, with follow-up beginning at transplantation and censored at 365 days. Schoenfeld-residual diagnostics indicated nonproportional hazards for the sex-by-era term and continuous PHM ratio; primary adjusted analyses therefore used Kaplan-Meier pseudo-observations at 365 days with identity-link linear regression and robust standard errors clustered by transplant center. The model included recipient sex, allocation era, their interaction, recipient age, race/ethnicity, BMI, diagnosis, ECMO, intra-aortic balloon pump, durable VAD/TAH support, donor age, ischemic time, and continuous donor-recipient PHM ratio. Severe PHM undersizing was evaluated in a parallel fixed-time model, with absolute 365-day mortality differences reported. Cox models and proportional-hazards diagnostics were retained as sensitivity analyses.

### Statistical Analysis

Continuous variables are reported as median (IQR) unless otherwise specified, and categorical variables as count (percentage). Absolute female-minus-male differences were reported alongside relative effect estimates. Transplantation and first actual offer were analyzed with cumulative incidence functions and Fine-Gray models. Conditional final acceptance was analyzed with binomial-logit models incorporating listing-center fixed effects and standard errors clustered by listing episode and donor. Final-graft PHM undersizing, the donor-characteristic/logistics composite, and treated rejection were analyzed with logistic or mixed-effects logistic regression; the rejection model included a center random intercept. Survival was analyzed with Kaplan-Meier estimates and 365-day pseudo-observation regression with center-clustered standard errors; Cox models were sensitivity analyses. Analyses used model-specific complete cases. Sex-by-era interactions assessed whether associations differed across allocation eras; null interactions were not interpreted as equivalence. All tests were 2-sided, with P<.05. Analyses were performed in R; full specifications are provided in the Supplemental Material.

## Results

### Study Population, Transplant Access, and First Offer

The transplant cohort included 38,417 recipients; women represented approximately one-quarter of recipients in both eras. Compared with men, women were younger, had lower BMI, were more often Black non-Hispanic or transplanted for congenital heart disease, and less often received durable VAD/TAH support. Donor and transplant characteristics also differed by recipient sex, particularly donor sex and transport distance. Analytic populations and endpoint-specific denominators are summarized in Supplemental Table S1.

Transplant access increased markedly after the allocation revision in both sexes. At 1 year after listing, the cumulative incidence of transplantation increased from 61.6% to 76.3% among women and from 55.4% to 71.4% among men. The female-minus-male difference was 6.2 percentage points pre-policy and 4.8 percentage points post-policy; the across-era change was −1.4 percentage points (95% CI, −3.4 to 0.7; P=.167), and the Fine-Gray sex-by-era interaction was not supported (sHR, 0.965; 95% CI, 0.921-1.011; P=.133). First actual offers also occurred earlier after the policy revision. The 1-year female-minus-male difference narrowed from −1.8 percentage points pre-policy to −0.5 percentage points post-policy, an across-era change of +1.3 percentage points (95% CI, 0.4-2.3). Although the interaction was statistically significant (adjusted sHR, 1.054; 95% CI, 1.011-1.099; P=.013), the absolute change was small. Full estimates are reported in Supplemental Table S2 and Supplemental Figure S2.

### Conditional Final Acceptance of Donor-Heart Offers

Offers made to female candidates had higher adjusted conditional final-acceptance probabilities across the offer sequence in both eras. In the primary model limited to the first 50 offers, the male-versus-female OR was 0.608 (95% CI, 0.540-0.684) pre-policy and 0.653 (95% CI, 0.581-0.734) post-policy. The sex-by-era interaction was not supported (OR, 1.074; 95% CI, 0.969-1.190; P=.172), and additional adjustment for donor characteristics produced a similar estimate (OR, 1.066; 95% CI, 0.957-1.188; P=.246). Excluding provisional Z responses did not materially change the findings. In the post-policy cohort, the association persisted after cPRA adjustment (male vs female OR, 0.607; 95% CI, 0.523-0.704; P<.001). Offer-specific and cumulative acceptance probabilities are reported in Supplemental Tables S3-S4 and Supplemental Figure S3.

### PHM Matching Across Offer and Transplant Stages

Crude severe PHM undersizing among actual offers was substantially less frequent for women than men (8.3% vs 44.3% pre-policy; 7.1% vs 40.9% post-policy), although recipient-PHM distributions differed markedly by sex. After standardizing women and men to the same pooled recipient-PHM and measured covariate distribution, severe-undersizing probabilities remained lower for women: 28.9% versus 38.3% pre-policy (female-minus-male difference, −9.4 percentage points; 95% CI, −13.1 to −5.9) and 25.5% versus 35.4% post-policy (difference, −9.8 percentage points; 95% CI, −13.5 to −6.3). The adjusted male-versus-female OR was 1.655 (95% CI, 1.484-1.846) pre-policy and 1.738 (95% CI, 1.637-1.844) post-policy; the sex-by-era interaction was not supported (P=.492). Findings were consistent within the common-support range. At the same standardized recipient PHM, offers made to women contained slightly greater donor PHM than offers made to men in both eras (male-minus-female difference, −5.2 pre-policy and −5.4 post-policy; both P<.001; interaction P=.699) (Figure 1A; Supplemental Tables S6-S9 and Supplemental Figure S4).

**Figure 1.**
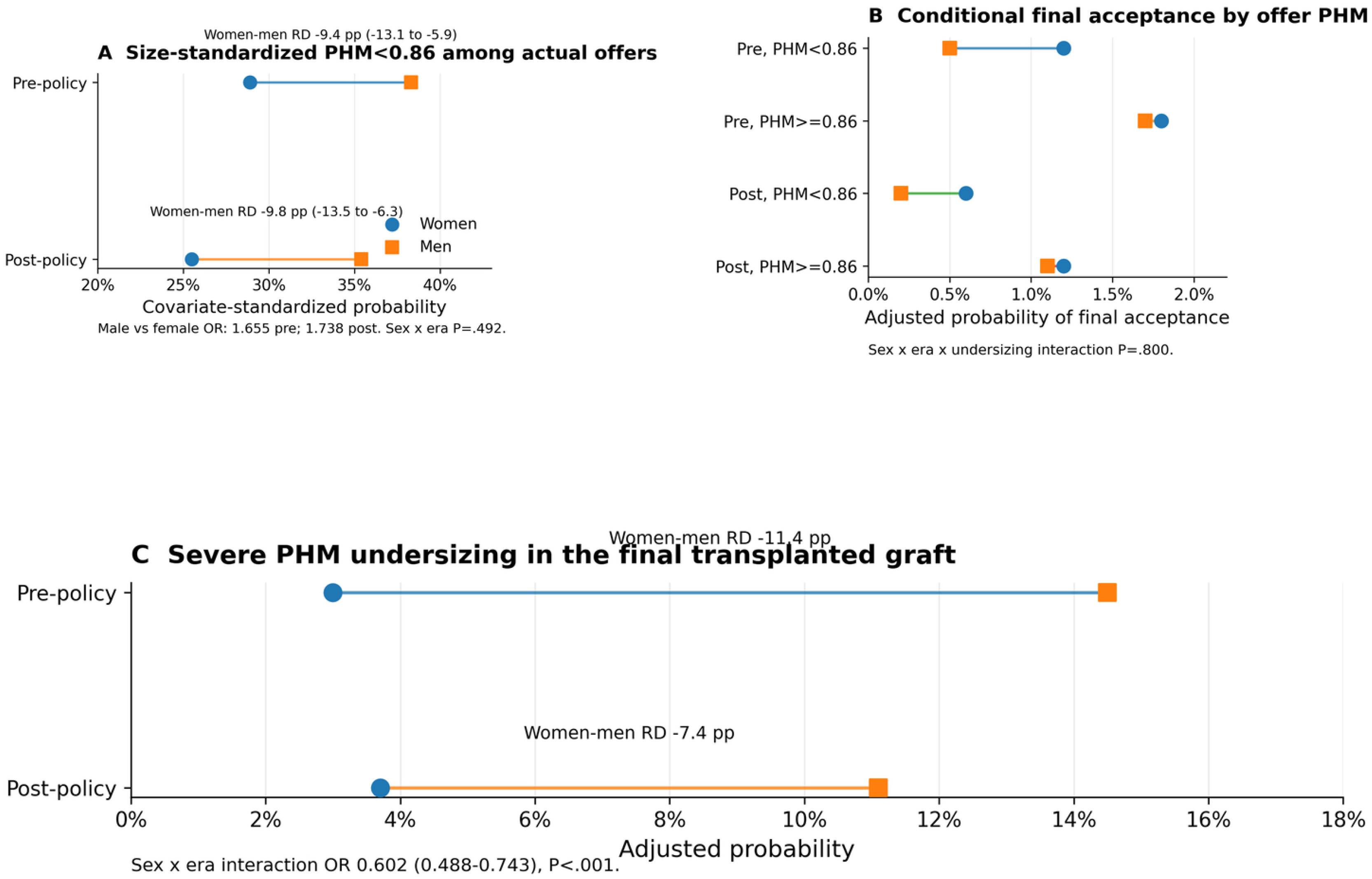
Sex differences in predicted heart mass matching across donor-heart offer exposure, conditional final acceptance, and transplantation. Panel A shows covariate-standardized probabilities of encountering an actual donor-heart offer with donor/recipient PHM ratio <0.86 after standardizing women and men to the same pooled recipient-PHM and measured covariate distribution. Panel B shows adjusted conditional final-acceptance probabilities for undersized and nonundersized offers. Panel C shows adjusted probabilities of severe PHM undersizing among transplanted grafts. Together, the panels show where the PHM difference was observed across the pathway; they do not identify a causal allocation mechanism.

Among offers with PHM ratio <0.86, adjusted final-acceptance probabilities were 1.2% for offers made to women versus 0.5% for offers made to men pre-policy and 0.6% versus 0.2% post-policy. The sex-by-era-by-undersizing interaction was not supported (P=.800) (Figure 1B; Supplemental Table S10).

The same direction was observed for final grafts. Severe PHM undersizing occurred in 4.0% of women versus 17.8% of men pre-policy and 4.8% versus 14.2% post-policy. Center-adjusted probabilities were 3.0% versus 14.5% pre-policy and 3.7% versus 11.1% post-policy. The sex difference was smaller post-policy (sex-by-era interaction OR, 0.602; 95% CI, 0.488-0.743; P<.001), but severe undersizing remained substantially less frequent among women in both eras (Figure 1C; Supplemental Table S10).

### Conventional Donor Profiles

Differences in conventional donor characteristics were generally small and inconsistent (Table 1). Pre-policy, women more often received donors aged >=50 years (9.3% vs 8.1%) and donors with diabetes (4.4% vs 3.4%). Post-policy, most biologic donor characteristics were similar by recipient sex. Ischemic time >=4 hours increased substantially in both sexes and was similar post-policy (32.0% among women vs 33.1% among men). Donor substance history was more common among male recipients in both eras.

**Table 1.** Conventional donor characteristics and final-graft predicted heart mass matching by recipient sex and allocation era.

| Characteristic | Pre women<br>(n=3,959) | Pre men<br>(n=11,251) | Post women<br>(n=6,159) | Post men<br>(n=17,048) |
| --- | --- | --- | --- | --- |
| Donor age $\geq 50$ y | 368 (9.3) | 915 (8.1) | 421 (6.8) | 1,141 (6.7) |
| Donor age $\geq 55$ y | 132 (3.3) | 316 (2.8) | 118 (1.9) | 388 (2.3) |
| Donor LVEF $< 50\%$ | 58 (1.5) | 165 (1.5) | 79 (1.3) | 246 (1.4) |
| Donor cardiac arrest/CPR | 277 (7.0) | 849 (7.6) | 356 (6.4) | 928 (6.2) |
| Ischemic time $\geq 4$ h | 726 (18.4) | 1,955 (17.4) | 1,919 (32.0) | 5,491 (33.1) |
| Donor diabetes | 175 (4.4) | 383 (3.4) | 255 (4.2) | 778 (4.6) |
| Donor substance history | 2,134 (54.1) | 7,172 (64.0) | 3,190 (52.9) | 9,725 (58.3) |
| Threshold donor-characteristic/logistics composite* | 1,229 (31.3) | 3,293 (29.5) | 2,456 (42.2) | 7,030 (44.2) |
| Final-graft PHM ratio, median [IQR] | 1.122 [0.996-1.278] | 0.976 [0.890-1.074] | 1.096 [0.984-1.238] | 0.982 [0.904-1.085] |
| Final-graft PHM $< 0.86$ | 158 (4.0) | 1,999 (17.8) | 296 (4.8) | 2,412 (14.2) |

\*Composite includes donor age $\geq 55$ years, donor LVEF $< 50\%$ , ischemic time $\geq 4$ hours, donor diabetes, or donor cardiac arrest/CPR. It is reported as a descriptive donor-characteristic/logistics summary and not as a unitary measure of graft quality.

**Table 2.** Key adjusted analyses from offer exposure through post-transplant outcomes.

| Domain | Key comparison | Effect estimate | Absolute/adjusted result | Interpretation |
| --- | --- | --- | --- | --- |
| First actual offer | Sex x era | sHR 1.054 (1.011-1.099); P=.013 | 1-y female-male difference: -1.8 pp pre; -0.5 pp post; across-era change +1.3 pp | Small attenuation of pre-existing first-offer difference |
| Conditional final acceptance | Sex x era | OR 1.074 (0.969-1.190); P=.172 | Male vs female OR: 0.608 pre; 0.653 post | Higher conditional final-acceptance probability for offers to women in both eras; no differential era change |
| Offer PHM $< 0.86$, size-standardized | Male vs female; sex x era | OR 1.655 pre; 1.738 post; interaction P=.492 | 28.9% vs 38.3% pre; 25.5% vs 35.4% post (women vs men) | Residual sex difference persists after recipient-size standardization |
| Final-graft PHM $< 0.86$ | Sex x era | OR 0.602 (0.488-0.743); P<.001 | Adjusted 3.0% vs 14.5% pre; 3.7% vs 11.1% post (women vs men) | Women less often receive severely undersized grafts in both eras |
| Threshold donor composite | Sex x era | OR 1.209 (1.091-1.340); P<.001 | 31.3% vs 29.5% pre; 42.2% vs 44.2% post (women vs men) | Small direction reversal; definition-sensitive |
| Treated rejection $\leq 1$ y | Male vs female; sex x era | OR 0.667 (0.589-0.756); P<.001; interaction P=.550 | Adjusted 22.5% vs 16.3% pre; 16.8% vs 11.4% post (women vs men) | Female excess persists; no differential era change |
| Post-policy treated rejection + cPRA | Male vs female | OR 0.678 (0.583-0.790); P<.001 | Adjusted 16.6% vs 12.0% (women vs men); RD 4.6 pp | cPRA attenuates but does not eliminate association |
| 365-day survival | Sex x era absolute change | -0.64 pp (-1.97 to 0.68); P=.341 | Adjusted survival: 90.2% vs 90.9% pre; 91.0% vs 92.3% post (women vs men) | No evidence of differential era change; post-policy female-male difference -1.29 pp |
| PHM $< 0.86$ and 365-day mortality | Pooled PHM mortality RD; PHM x era change | +1.43 pp (0.33 to 2.54); P=.011; across-era change -0.76 pp (-2.96 to 1.45); P=.502 | Adjusted mortality RD: +1.81 pp pre; +1.06 pp post | Modest absolute mortality difference; no evidence of era modification |

Abbreviations: cPRA, calculated panel-reactive antibody; OR, odds ratio; PHM, predicted heart mass; pp, percentage points; RD, risk difference; sHR, subdistribution hazard ratio.

The threshold donor-characteristic/logistics composite was more frequent among women pre-policy (31.3% vs 29.5%) and among men post-policy (42.2% among women vs 44.2% among men). The center-accounted sex-by-era interaction OR was 1.209 (95% CI, 1.091-1.340; P<.001). Alternative composite definitions changed both the direction and magnitude of the sex differences. Accordingly, the composite was interpreted as descriptive donor-profile information rather than a unitary measure of graft quality. Definitions, sensitivity analyses, and center effects are reported in Supplemental Table S11.

### Registry-Reported Treated Rejection

Among 34,408 recipients eligible for 1-year rejection assessment, TRTREJ1Y was reported as yes or no in 29,958 (87.1%); 27,426 complete cases entered the adjusted model. Registry-reported treated rejection was more frequent among women. In the center-random-intercept model, the male-versus-female OR was 0.667 (95% CI, 0.589-0.756; P<.001). Adjusted probabilities were 22.5% for women versus 16.3% for men pre-policy and 16.8% versus 11.4% post-policy. The female-minus-male difference was 6.2 percentage points pre-policy and 5.3 percentage points post-policy, with no supported sex-by-era interaction (OR, 0.957; 95% CI, 0.829-1.105; P=.550). Center-level heterogeneity remained measurable (ICC, 0.118) (Figure 2A; Supplemental Table S12).

**Figure 2.**
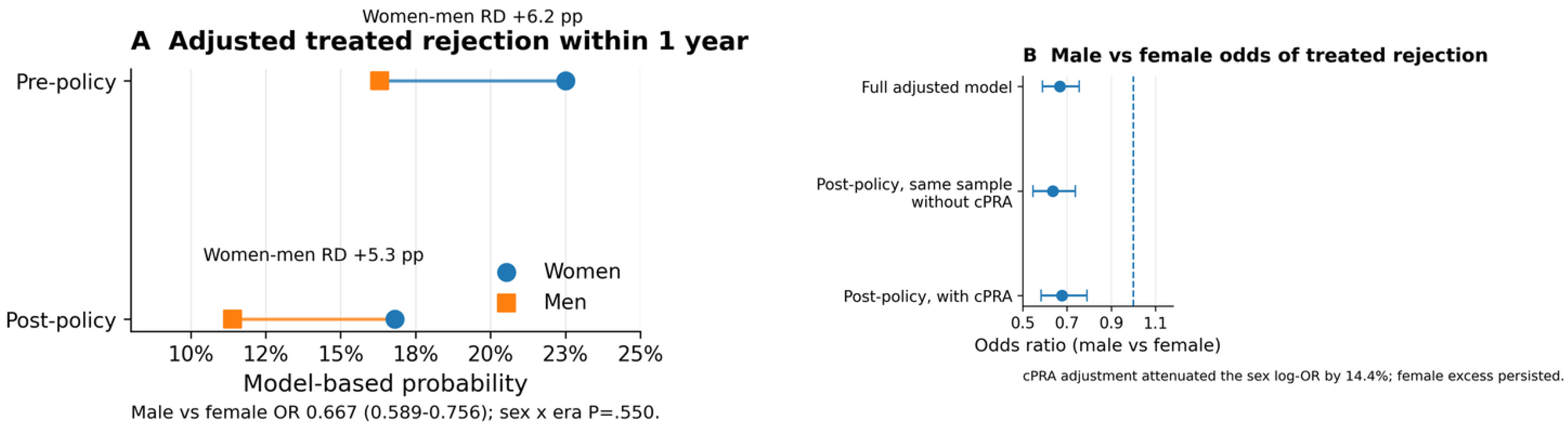
Adjusted registry-reported treated rejection by recipient sex and allocation era. Panel A shows model-based probabilities of treated rejection within 1 year among recipients with at least 1 year of potential follow-up after adjustment for recipient, support, donor, PHM, and transplant-center factors. Panel B shows male-versus-female adjusted odds ratios with 95% confidence intervals for the full pre/post model and post-policy same-sample models before and after cPRA adjustment. cPRA adjustment attenuated the sex log-odds coefficient by 14.4%, but the association persisted.

Pretransplant cPRA could be reconstructed reliably from waitlist history only in the post-policy era (92.7% complete among women and 92.8% among men). In a contemporary same-sample sensitivity analysis, adding cPRA attenuated the sex log-OR by 14.4%, but the association persisted (male vs female OR, 0.678; 95% CI, 0.583-0.790; P<.001). After cPRA adjustment, estimated treated-rejection probabilities were 16.6% for women and 12.0% for men (difference, 4.6 percentage points; 95% CI, 2.7-6.6) (Figure 2B; Supplemental Table S13).

### Post-Transplant Survival and PHM-Associated Mortality

Unadjusted Kaplan-Meier survival differed statistically across the four sex-by-era groups, although absolute differences were modest. Survival at 1, 3, and 5 years was 91.3%, 86.3%, and 80.9% among pre-policy women; 90.8%, 84.1%, and 77.4% among post-policy women; 91.2%, 85.6%, and 80.3% among pre-policy men; and 91.4%, 84.6%, and 78.1% among post-policy men. In the adjusted pseudo-observation analysis (N=34,273; 2,937 deaths), standardized 365-day survival was 90.2% (95% CI, 89.2%-91.2%) among pre-policy women, 90.9% (95% CI, 90.3%-91.5%) among pre-policy men, 91.0% (95% CI, 90.1%-91.9%) among post-policy women, and 92.3% (95% CI, 91.7%-92.8%) among post-policy men. The female-minus-male difference was −0.64 percentage points pre-policy (95% CI, −1.63 to 0.35; P=.202) and −1.29 percentage points post-policy (95% CI, −2.16 to −0.42; P=.004); the across-era change was −0.64 percentage points (95% CI, −1.97 to 0.68; P=.341) (Figure 3A-B; Supplemental Table S14).

**Figure 3.**
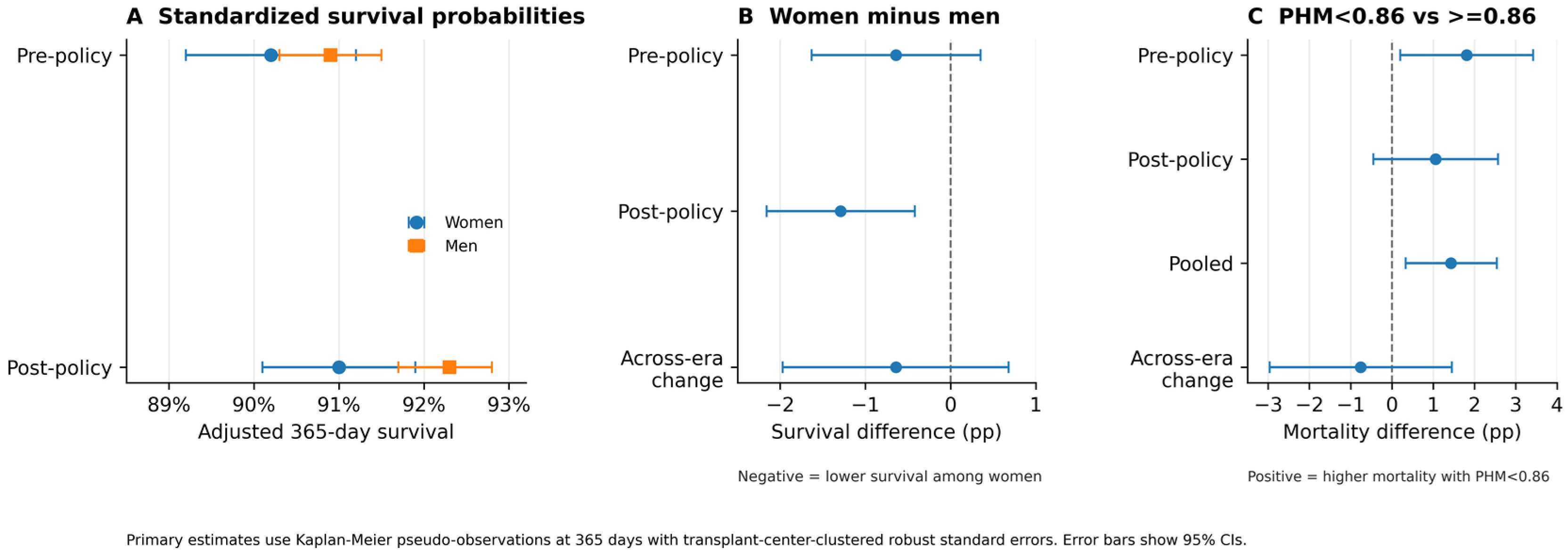
Adjusted 365-day post-transplant survival and absolute mortality differences. Panel A shows covariate-standardized 365-day survival probabilities with 95% confidence intervals by recipient sex and allocation era. Panel B shows female-minus-male absolute survival differences within each era and the change in that difference across eras; negative values indicate lower survival among women. Panel C shows adjusted absolute mortality-risk differences associated with severe PHM undersizing within each era, the pooled difference, and the across-era change; positive values indicate higher mortality with PHM <0.86. Primary estimates were obtained from Kaplan-Meier pseudo-observation regression with robust standard errors clustered by transplant center; follow-up was censored at 365 days.

In the PHM clinical-validation cohort (N=37,660; 3,141 deaths), severe PHM undersizing was associated with a pooled adjusted absolute increase in 365-day mortality of 1.43 percentage points (95% CI, 0.33-2.54; P=.011). The adjusted mortality difference was 1.81 percentage points pre-policy (95% CI, 0.20-3.42; P=.027) and 1.06 percentage points post-policy (95% CI, −0.45 to 2.57; P=.170), with no supported across-era change (−0.76 percentage points; 95% CI, −2.96 to 1.45; P=.502) (Figure 3C; Supplemental Table S14).

## Discussion

In this linked national STAR-PTR analysis, associations with recipient sex were stage-specific and did not align along a single axis of advantage or disadvantage. Offers made to female candidates had higher adjusted conditional final-acceptance probabilities in both eras. After standardization to a common recipient-size and covariate distribution, women encountered fewer PHM-undersized offers and received fewer undersized grafts. In contrast, conventional donor profiles were heterogeneous, women had more adjusted registry-reported treated rejection, and 365-day survival was modestly lower among women post-policy without a significant across-era change. These findings show why offer exposure, acceptance, matching, and post-transplant outcomes should be evaluated separately.

The offer-level findings extend Breathett et al., who found higher early conditional acceptance and cumulative final acceptance for offers made to women after 2018. (14) We observed the same overall direction in both allocation eras, without evidence that the sex association with conditional final acceptance changed materially after 2018. Our linked analysis further separated realized offer composition from the subsequent acceptance decision. The sex difference in severe PHM undersizing was detectable among actual offers after recipient-size standardization, whereas conditional final acceptance of an undersized offer was not lower for female candidates. Thus, the final-graft PHM difference was not attributable solely to preferential decline of undersized offers for women. The data cannot identify which allocation, geographic, center-filtering, or clinical processes generated the realized offer distribution.

PHM provides a clinically interpretable framework for this stage-localization analysis, although its prognostic importance is context dependent. Kransdorf et al. showed that PHM predicted 1-year mortality more effectively than weight, height, body mass index, or body surface area and identified increased mortality below a donor-to-recipient PHM ratio of approximately 0.86. (17) Tao et al. subsequently reported higher mortality with PHM undermatching in selected recipients, including those with elevated pulmonary vascular resistance. (19) By contrast, DeFilippis et al. found that low PHM ratios were associated with 1-year mortality in a historical era but not under the contemporary allocation system. (18) In our cohort, PHM <0.86 was associated with a modest pooled absolute increase in 365-day mortality, without evidence of era modification. Recipient size explained much of the crude contrast, but a residual 9- to 10-percentage-point difference remained after standardization and within common support. PHM should therefore be interpreted as a specific matching measure, not a unitary marker of donor-heart quality or a purely mathematical proxy for male recipient size.

The conventional donor-profile findings provide complementary context. Jain et al. emphasized that donor age, ventricular function, ischemic time, and other commonly labeled high-risk features have effects that depend on recipient selection, preservation, and logistics. (20) Consistent with that framework, our threshold composite showed small, definition-sensitive differences that reversed direction across eras. Moayedi et al. likewise reported that women who reached transplantation had fewer recipient risk factors but received donors with higher measured risk profiles; after matching recipient and donor characteristics, survival did not differ by recipient sex. (21) Conventional donor features describe the utilized donor pool, but their sum is not a validated scalar measure of graft quality or inequitable allocation.

The persistent excess of registry-reported treated rejection among women was a clinically distinct downstream finding. Kim et al., in the 2026 ISHLT Registry report, found greater allosensitization among female recipients. (4) Nguyen et al., using biopsy-proven antibody-mediated rejection, found that female recipient sex remained independently associated with rejection after adjustment for preformed donor-specific antibodies, HLA mismatch, size mismatch, and other factors. (25) Perry and Sen described plausible contributions from pregnancy-associated alloimmunization, sex-chromosome and hormonal effects, and differential immunosuppressant metabolism. (24) In our post-policy sensitivity analysis, cPRA adjustment attenuated the sex coefficient by 14.4%, consistent with sensitization contributing to, but not fully accounting for, the association. This was not a mediation estimate, and the UNOS endpoint does not distinguish cellular from antibody-mediated rejection or capture donor-specific antibodies, surveillance, or treatment thresholds. Biological and care-delivery explanations therefore remain unresolved.

Survival findings in the literature vary by cohort, donor-recipient pairing, follow-up horizon, and estimand. Moayedi et al. found no significant survival difference after matching women and men on recipient and donor risk. (21) Vinson et al., in an individual-patient-data meta-analysis of more than 100,000 recipients, reported higher excess mortality among female recipients across several donor-sex and age strata, particularly with male donors. (22) Purohit et al. reported lower adjusted 1-year survival among women under both previous and revised U.S. systems, and Ramu et al. found a 14% higher adjusted mortality hazard among women at both 1 and 5 years after 2018. (12,13) Conversely, Kim et al. reported similar 1-year survival by recipient sex within contemporary ISHLT eras. (4) In our fixed-time analysis, adjusted 365-day survival was modestly lower among women post-policy, but the female-male difference did not change significantly across eras. This null interaction is not evidence of equivalence; it indicates no supported differential era change in short-term survival.

The allocation-era comparison is best understood as temporal context rather than a causal policy experiment. DeFilippis et al. documented increased transplantation opportunities for women during the first 2 years after the 2018 revision. (11) Purohit et al. later found higher 1-year transplantation incidence among women in both systems, while Ramu et al. reported higher transplantation and lower delisting for death or deterioration among women under the revised system. (12,13) In our cohort, access increased for both sexes, the small first-offer difference narrowed, and most sex-by-era interactions across acceptance, PHM exposure, rejection, and survival were not supported. These findings do not isolate a policy effect; they show that several stage-specific associations were observed under two substantially different allocation systems.

Several limitations should guide interpretation. STAR-PTR linkage captures realized offers and cannot disentangle allocation rules from geography, candidate filters, center practice, or unmeasured clinical judgment. PTR capture, donor linkage, and offer-level PHM completeness were high but imperfect. Despite size standardization and common-support restriction, residual confounding remains possible, and PHM does not capture all relevant cardiac geometry or recipient physiology. The threshold donor composite was not externally validated as a graft-quality score.

Outcome analyses have additional limitations. cPRA was sufficiently complete for adjustment only post-policy. Treated rejection was a follow-up-derived binary endpoint without exact event dates; unknown or missing status was excluded, death could truncate ascertainment, and rejection phenotype, donor-specific antibody timing, biopsy protocols, and treatment thresholds were unavailable. Primary graft dysfunction was too incompletely recorded for reliable era comparison. Survival analyses used complete cases and fixed-time pseudo-observation regression because proportional-hazards diagnostics were significant for key Cox terms. Finally, registry-recorded sex does not capture gender identity, and null interaction tests indicate absence of evidence for differential era modification rather than proof of no difference.

In conclusion, recipient-sex associations differed across the heart-transplant pathway rather than forming a uniform gradient. The PHM difference was detectable among realized offers after recipient-size standardization and was not attributable solely to lower conditional final acceptance of undersized offers for female candidates. Conventional donor profiles, treated rejection, and short-term survival followed different directions. Transplantation performance should assess offer exposure, conditional acceptance, matching, and downstream outcomes separately before inferring pathway-wide advantage or disadvantage from one measure.

## Declarations

### Ethics approval

This study used deidentified United Network for Organ Sharing registry data and was determined exempt by the Institutional Review Board (IRB No. 172, Pro00073879; approved May 29, 2016). Informed consent was waived.

### Data availability statement

The data that support the findings of this study were provided by the United Network for Organ Sharing under a data use agreement. Deidentified registry and match-run data are available to qualified investigators from UNOS/OPTN and cannot be redistributed by the authors.

### Author contributions

J.S.M. and Y.D.B. conceived and designed the study. J.S.M. performed data curation and statistical analysis and drafted the manuscript. O.G., A.L.L., E.P., M.S., and E.H.K. contributed to interpretation and critical revision. All authors approved the final version and agree to be accountable for the work.

## Supporting Information

Supporting information includes supplemental methods; analytic cohort and endpoint-completeness tables; detailed first-offer and conditional-acceptance estimates; offer-capture and external cohort QA; PHM size-standardization, common-support, and offer-to-transplant analyses; donor-composite sensitivity results; treated-rejection and cPRA models; mortality analyses; variable-availability audits; exact model specifications; and five supplemental figures.

